# Physical activity bout length and risk of major adverse cardiovascular events in adults with hypertension

**DOI:** 10.1101/2024.07.31.24311326

**Authors:** Matthew N. Ahmadi, Angelo Sabag, Raaj Biswas, Borja del Pozo Cruz, Clara K. Chow, Emmanuel Stamatakis

## Abstract

**Background:** Hypertension is a major risk factor for cardiovascular disease. Although it is well established that physical activity is cardioprotective, it is less clear how cardiovascular stress-related properties (i.e. intensity and bout length) determine future cardiovascular risk in adults with hypertension.

**Objectives:** We examined the dose-response associations of moderate and vigorous physical activity bouts of variable length with major adverse cardiovascular events (MACE) and its sub-types (stroke, myocardial infarction, heart failure) in adults with hypertension.

**Methods:** Participants of the UK Biobank wearables sub-study with a clinical diagnosis of hypertension. Short bouts of moderate activity were classified as lasting up to 3 minutes and for vigorous activity up to 1 minute. Long bouts of moderate activity were classified as lasting >5 minutes, and for vigorous activity >2 minutes. In sensitivity analyses we also tested alternative vigorous intensity bout length definitions for short (up to 2 minutes) and long (> 3, >4, and >5 minutes).

**Results:** Among 36,957 participants (62.1 (SD= 7.7) years; 58.4% female) with an average follow up of 7.9 (1.1) years, 1,374 MACE, 394 stroke, 623 myocardial infarction, and 357 heart failure events occurred. Moderate intensity was associated with lower risk of MACE and its individual sub-types for both short (<3 mins) and long (>5 mins) bout lengths with a stronger dose-response magnitude for longer bouts. We observed a consistent inverse dose response association for vigorous intensity accrued through short bouts (<1 min) and overall MACE, stroke, myocardial infarction, and heart failure risk. The median duration of 3.5 minutes/day corresponded to a hazard ratio (HR) ranging between 0.57 [0.39, 0.83] for heart failure to 0.66 [0.46, 0.96] for stroke.

In contrast, vigorous intensity activity accrued through longer bouts showed a steep linear dose-response association for higher stroke risk. The median (6.0 minutes/day) and maximum (9.0 minutes/day) of activity accrued through vigorous bouts lasting >2 mins were associated with stroke HRs of 2.06 [1.38, 3.07] and 2.80 [1.72, 4.56], respectively. Additional analyses in 1 extra minute bout length increments revealed that the higher the “long bout” threshold the steeper the stroke risk, e.g the median of bouts lasting >5 mins (6.4 minutes/day) was associated with a HR of 2.69 [1.72, 4.21].

**Conclusion:** For adults with hypertension, moderate intensity and short bouts of vigorous intensity were beneficially associated with lower MACE, stroke, myocardial infarction, and heart failure risk. Vigorous intensity accumulated in long bouts showed a steep deleterious dose-response with stroke risk, and were not associated with lower overall MACE, myocardial infarction, or heart failure. Our results highlight the importance of bout length for vigorous intensity as a determining factor for cardiovascular health in adults with hypertension.

## Introduction

Cardiovascular disease (CVD) is the leading cause of death worldwide with prevention strategies focused on mitigating cardiometabolic and lifestyle risk factors^12^. Hypertension is the leading cardiometabolic contributor to CVD, particularly cerebrovascular events, with an estimated one in three adults living with the condition of which only 20% have controlled hypertension^3–5^. While the cardioprotective benefits of physical activity are well-established, some research suggests that high intensity physical activity may lead to an increased risk of cardiovascular events and this may be more apparent in adults that are less fit or have pre-existing cardiovascular conditions, including hypertension^6–8^. Specifically, in people with hypertension, physical activity of higher intensity may provide no clear protective effects^9–11^ or even increase stroke risk^12–14^.

In light of this evidence, the American Heart Association and European Society of Cardiology clinical guidelines precautionary advise against prescribing high-intensity physical activities to people with hypertension ^13^ ^15–18^. Such guidance has been informed predominantly by studies that used self-report questionnaires to quantify the duration and intensity of physical activity, which cannot capture activity bouts lasting less than 10-15 minutes. In addition, it is based on studies that evaluated total volume of high intensity activities irrespective of how it was accumulated (i.e. in short intermittent bursts vs prolonged sustained bouts.)

Recent wearable device-based studies have significantly enhanced our understanding of how short and long activity bouts impact cardiovascular health in healthy adults. Short bouts of vigorous intensity lasting up to 1 minute may confer measurable health benefits and reduce the risk of CVD^19^ ^20^. Other studies have indicated short bursts of moderate to vigorous physical activity lasting only a few minutes (eg. 1-5 minutes) have similar associations with favourable cardiovascular outcomes as prolonged bouts lasting several minutes (>10 minutes)^21–23^. Clinical trials in adults with cardiovascular morbidities have shown short bouts and low volumes of high intensity activity can positively affect cardiometabolic markers. This includes improved lipid profile, glucose regulation, and reduced arterial stiffness^24^ ^25^. However, a significant gap remains in our understanding of how activity bout length may influence long-term risks and benefits for major adverse cardiovascular events (MACE), in adults with hypertension.

We examined the dose-response associations of wearable device-based moderate and vigorous physical activity volume accrued through short and long bouts with MACE risk and its sub-types (stroke, myocardial infarction, heart failure).

## Methods

### Sample

Participants were included from the UK Biobank Study. All participants were enrolled between 2006-2010 and provided informed written consent. Ethical approval was provided by the UK’s National Health Service, National Research Ethics Service (Ref 11/NW/0382). Participants completed physical examinations by trained staff and touchscreen questionnaires. We defined clinically diagnosed hypertension using 1) hospitalisation admission records; 2) general practitioner records and self-report data; 3) as well as those taking hypertensive medication using linked general practitioner medication records. Prescribed hypertensive medications included ACE inhibitors, Angiotensin II receptor blockers, calcium channel blockers, beta-blockers, diuretics (eg. bendroflumethiazide and chlortalidone), alpha blockers, renin inhibitors, central agonists, peripheral adrenergic inhibitors, and vasodilators. **Supplemental Table 1** provides a complete list of ICD-10, Read Codes, and BNF Codes used to identify participants. We excluded participants with missing data, prevalent CVD (ascertained through self-report and hospital admission records), and an event within the first 12 months of follow up^20^ ^21^ ^26^ (**Figure 1)**.

### Physical activity assessment

Between 2013 and 2015, 103,684 participants wore an Axivity AX3 accelerometer (Axivity Ltd, Newcastle upon Tyne, UK) on their dominant wrist for 24-hrs/day for 7 days. Devices were calibrated and non-wear periods were detected using standard procedures^27^. We included participants with ≥3 valid wear day (≥16 hours of wear-time) with at least one of those days being a weekend day. Physical activity intensity was classified with a validated accelerometer-based two-level Random Forest^28^ machine learning activity scheme that has previously been used in the UK Biobank cohort^19^ ^21^. Briefly, this activity scheme uses features in the raw acceleration signal to identify different activity types and then quantify time spent in different activity intensities in 10 second windows. To calculate physical activity duration across all valid wear days, we summed time spent in each respective activity intensity band as light, moderate, and vigorous. For moderate intensity, we calculated the duration from short bouts lasting up to 3 minutes, and long bouts lasting >5 minutes. We calculated vigorous intensity duration from short bouts lasting up to 1 minute and long bouts lasting >2 minutes. Our bout length thresholds were based on activity accumulation patterns where 90.8% of moderate intensity occurred in up to 3 minute bouts, and 90.5% of all vigorous intensity occurred in up to 1 minute bouts and prior research showing health benefits from bout thresholds^19–21^.

### Major adverse cardiovascular event ascertainment

Participants were followed up through November 30^th^, 2022, with deaths obtained through linkage with the National Health Service (NHS) Digital of England and Wales or the NHS Central Register and National Records of Scotland. Inpatient hospitalisation data were provided by either the Hospital Episode Statistics for England (October 31^st^, 2022), the Patient Episode Database for Wales (May 31^st^, 2022), or the Scottish Morbidity Record for Scotland (August 31^st^, 2022). MACE was defined as fatal cardiovascular events, or incidence of ST-elevation or non-ST elevation myocardial infarction, stroke, and heart failure. **Supplemental Table 2** provides the full list of specific ICD-10 codes.

### Covariates

Based on a directed acyclic graph (**Supplemental Figure 2)** our covariates, measured during clinic visits, included: age, sex, smoking status, alcohol consumption, fruit and vegetable consumption, sedentary time, sleep duration, ambulatory light intensity activity, highest attained education level, frailty index score^29^, medication use (diabetes, hypertension, high cholesterol), prevalent cancer, and family history of CVD and cancer. We also adjusted for moderate and vigorous intensity activity coming from bouts that were not part of the exposure. For example, in our analysis of vigorous intensity duration from short bouts lasting up to 1 minute, we included as a covariate the duration of vigorous intensity from bouts lasting >1 minute and total moderate intensity duration. In sensitivity analyses, we included as confounders clinical factors that may be mediators: waist circumference, glycated hemoglobin A1C, high-density and low-density lipoprotein, and triglycerides. Complete covariate definitions are provided in **Supplemental Table 3. Analyses**

We used Cox proportional hazards regression models to estimate hazard ratios (HR) with 95% CIs. Fine-Gray subdistribution was used for MACE and sub-type analyses with non-outcome deaths treated as a competing risk. We examined the dose-response association of short and long moderate and vigorous intensity bouts with MACE and sub-type risk using restricted cubic splines with knots evenly spaced along the intensity duration distribution (eg. 10^th^, 33^rd^, and 67^th^ percentiles for right skewed data). Departure from linearity was assessed by a Wald test. We tested proportional hazards assumptions in Cox’s regression models using Schoenfeld Residuals and no violations were observed (all p>0.05). The reference group was zero minutes of the physical activity exposure. We further calculated the adjusted dose-response absolute risk for each exposure and cardiovascular event outcome using Poisson regression, with knots at the same placement as the primary analyses. To further assess the potential effects of short and long bout lengths with stroke risk among adults with hypertension, we carried out the following additional analyses: 1) the influence of incremental 1 minute increases in the length of short and long activity bouts; 2) restricting vigorous intensity analyses to: a) participants who exercised; b) exercisers with sedentary occupations; c) participants with manual labour occupations; 3) risk stratified by age for young (<60 yrs) and older (≥60 yrs) adults; 4) risk stratified by diagnosis type (hospitalisation; prescribed medication; GP diagnosis).

We calculated E-values to estimate the plausibility of bias from unmeasured confounding for the median daily duration point estimates. To further assess the influence of alternate short and long vigorous bout length thresholds, we categorised short bouts as up to 2 minutes, and long bouts as >3, >4, and >5 minutes for each outcome. We included sensitivity analyses for adjustments of clinical biomarkers that maybe potential mediators that include glycated hemoglobin (HbA1c), low-density lipoprotein (LDL), high-density lipoprotein (HDL), and triglycerides. We also provide adjustment for central adiposity (waist circumference; sex-specific categories). We conducted analyses to minimize bias attributable to reverse causation by excluding: 1) underweight participants (body mass index <18.5kg/m^2^) or participants with fair or poor self-rated health; 2) high frailty (score ≥3 on a 0-5 scale); 3) current smokers; 4) exclusion of the first 5 years of follow-up. To determine if total intensity duration can obscure the influence of bout length on the associations, we assessed the dose-response association for total duration of 1) moderate; 2) vigorous; 3) and moderate to vigorous physical activity with MACE and each sub-type.

We reported this study according to Strengthening the Reporting of Observational Studies in Epidemiology (STROBE) guidelines (see **STROBE Statement** in the Supplement). We performed all analysis using R statistical software version 4.3.1 with the RMS version 6.7.0 and survival package version 3.5.5.

### Results

36,957 participants were included in our main analyses, with an average age of 62.1 (SD=7.7) years, of whom 21,585 (58.4%) were females, and 23,082 (62.5%) had a high school or college/university degree. Participants were followed up for an average of 7.9 (1.1) years. There was a total of 1,374 MACE, 357 heart failure, 623 myocardial infarction, and 394 stroke events. Participants had a median 24.0 [14.5, 37.0] minutes/day of moderate intensity accrued from up to 3 minute bouts and 3.5 minutes/day of vigorous intensity accrued from up to 1 minute bouts. **Table 1** provides complete participant characteristics by vigorous intensity duration. **Supplemental Figure 3-6** show the adjusted absolute risk dose-response association per 1,000 person-years.

### Overall major adverse cardiovascular events

In our MACE analyses, moderate intensity activity accumulated from short bouts lasting up to 3 minutes and long bouts >5 minutes, we observed an L-shaped inverse association with MACE risk (**Figure 1A**). For example, the daily median duration (10 minutes/day) of long bouts had an HR of 0.79 [0.66, 0.95]. Daily vigorous intensity duration accumulated from short bout lengths lasting up to 1 minute showed a protective non-linear association (**Figure 1B**). The median duration of 3.5 minutes/day corresponded to an HR of 0.62 [0.51, 0.76] with continued lower risk for higher daily duration. For long vigorous intensity bout lengths lasting >2 minutes, we observed a non-significant (ie. lower 95% CI crossed HR=1) trend for higher MACE risk as daily duration increased. This association pattern was consistent for 1 minute increment increases for thresholds of long bouts ranging from >3 minutes to >5 minutes (**Supplemental Figure 7**).

### Stroke

Both short and long bouts of moderate intensity were associated with lower stroke risk **(Figure 2A**). A median duration of 10 minutes/day from long bouts had an HR of 0.77 [0.61, 0.96], and the association for short bouts became significant when the duration exceeded 28 minutes/day. We observed an inverse linear association between higher daily vigorous intensity duration and stroke risk from short bouts with the median 3.5 minutes/day associated with an HR of 0.65 [0.46, 0.94] **(Figure 2B**). For vigorous intensity accumulated from long bouts lasting >2 minutes, we observed a linear association for higher stroke risk (**Figure 2B**). At a median duration of 6 minutes/day the HR was 2.06 [1.38, 3.07] and a maximum duration of 9 minutes/day had an HR of 2.80 [1.72, 4.56].

We observed an attenuation of the protective dose-response association as the threshold for short bout length increased and a gradient for higher risk as the length of long bouts increased incrementally by 1 minute (**Figure 3**). A median and maximum dose of 6.4 and 8.7 minutes/day coming from long bouts lasting >5 minutes corresponded to HRs of 2.69 [1.72, 4.21] and 3.77 [1.62, 8.73], respectively. To better understand these divergent associations between short and long bout lengths on stroke risk, we repeated analyses among exercisers and stratified by occupation. For exercisers and sedentary occupations (e.g.: office workers), we observed a general consistency in the dose-response association of vigorous intensity bout length for magnitude and direction with our equivalent results in the core sample (**Supplemental Figure 8**). For manual labour occupations, the protective association for short bouts was consistent, but with heightened stroke risk for long bouts. There were wide 95% CI’s in these sub-group analyses due to the smaller sample size.

### Myocardial infarction

We observed lower myocardial infarction risk for higher daily moderate intensity activity accrued through long bouts, and no association for short bouts (**Figure 4A**). A median duration of 10 minutes/day from long bouts had an HR of 0.64 [0.48, 0.87]. For vigorous intensity, both short and long bout lengths were associated with lower myocardial infarction risk (**Figure 4B**). For short bouts, we observed an L-shaped association that levelled-off at about 4.5 minutes/day (HR= 0.63 [0.47, 0.84]) and an inverse linear association with wide 95% CI’s for long bouts. There were negligible differences in the association pattern for progressively longer bout thresholds (HR difference <0.02 between different long bout lengths in 1 minute increment increases; **Supplemental Figure 9**).

### Heart failure

For moderate intensity, we observed lower heart failure risk from both short and long bout lengths with a stronger magnitude of association for the long bout length. A median of 24 minutes/day from short bouts and 10 minutes/day from long bouts was associated with an HR of 0.49 [0.31, 0.78] and 0.52 [0.38, 0.72], respectively. We observed an inverse linear association between short vigorous intensity bouts and heart failure risk with a median 3.5 minutes/day associated with an HR of 0.57 [0.39, 0.83] **(Figure 5A**). We did not observe a clear association between long vigorous intensity bouts lasting >2 minutes (**Figure 5A**) or successively longer bouts of >3, >4, or >5 minutes (**Supplemental Figure 10**).

### Additional and sensitivity analyses

Our analyses adjusting for clinical biomarkers and waist circumference showed association patterns that were consistent with our main analyses (**Supplemental Figures 11-14)**. Associations remained generally consistent after excluding participants with: 1) a high frailty index (score ≥3 on a 0-5 scale), 2) self-rated poor health, or a body mass index <18.5 kg/m^2^; 3) current smokers; 4) an event within the first 5 years of follow-up (**Supplemental Figure 15-22**). Our E-values suggest a moderate to high degree of unmeasured confounding would be required to reduce our observed associations to null. For example, the minimal dose E-values ranged from 2.45 [lower 95% CI: 1.32] for myocardial infarction to 2.90 [1.70] for heart failure (**Supplemental Table 4**). Analysis of total moderate and moderate to vigorous physical activity duration showed a beneficial association with total MACE (and each sub-type) (**Supplemental Figure 23-26**). Total daily duration of vigorous intensity showed a beneficial association with MACE, myocardial infarction, and heart failure, but not stroke risk. Additional stroke analyses were overall consistent with the main results and showed higher risk from long vigorous bouts for older adults (≥60 yrs) with a steep linear increase in risk after about 5 minutes/day for young adults (<60 yrs) (**Supplemental Figure 27A**). Stratification by diagnosis type showed attenuated risk for long bouts among adults taking hypertensive medication (**Supplemental Figure 27B**). In all these additional stroke analyses 95% CI’s were wider due to the smaller sample size in the stratified analytic groups. Using an alternate short vigorous bout length threshold of “up to 2 minutes” showed consistent results as our main analysis for each outcome (**Supplemental Figure 28 and Figure 3**).

## Discussion

Using wearable device-based data and focused on adults with a clinical diagnosis of hypertension, our study is the first to investigate the potential role of bout length as a determinant in the effects of physical activity intensity on major adverse cardiovascular events. We found consistent beneficial dose-response associations (linear or near linear) for lower total MACE risk, as well as incident stroke, myocardial infarction, and heart failure from higher daily durations of moderate or vigorous intensity accumulated through short bouts. In contrast, there was no protective association for vigorous activity accrued through longer bouts. Notably a pronounced and steep linear association gradient for stroke events, that showed an approximate 2 to 3-fold higher risk from long bouts of vigorous intensity.

We found an inverse linear association between vigorous intensity and total MACE with a median 3.5 minutes/day corresponding to 38% lower risk with the potential for continued lower risk from higher daily durations. These potential cardioprotective benefits were accrued specifically through short bouts. These results are in sharp contrast to previous studies reliant on self-reported physical activity that found deleterious associations between vigorous intensity activity and cardiovascular events^10^ ^30^ ^31^. Notably, self-report physical activity is limited to only measuring prolonged and sustained bouts of activity. When we assessed longer bouts of vigorous intensity lasting >2 to >5 minutes, we did not observe a protective association. The findings of this study are consistent and extend on that of prior small randomised control trials and meta-analyses in hypertensive adults that found short 30 to 90 second intervals of high intensity activities can improve vasodilation, blood and plasma viscosity, and reduce fibrinogen concentration^32^ ^33^. Our findings suggest current CVD prevention strategies for adults with hypertension, which primarily recommend light or moderate intensity activities^8^, may overlook significant health benefits that can still be derived from short, but not long, bouts of vigorous intensity activity. These findings underscore the potential need for a reinterpretation of the evidence base.

### Stroke

The American Heart Association^7^, European Society of Cardiology^18^, and previous research^12^ ^34^ has suggested vigorous intensity activity can increase stroke risk, that is attributable to vascular stress and strain leading to deleterious changes in the cardiovascular structure, such as arterial stiffness, coronary artery calcification, diastolic dysfunction and cervical artery dissection^6^ ^10^ ^35^. Available evidence, however, has been limited to assessing total volume accrued primarily through long continuous activity bouts. Our study, using wearable devices to continuously record physical activity at a high resolution, highlights the importance of bout length in the vigorous intensity exposure. Examination of long activity bouts showed association patterns consistent with previous research where vigorous intensity was associated with higher risk and moderate intensity was associated with lower risk. When assessing specifically short activity bout lengths, we found vigorous intensity was associated with a 34-45% (median to maximum daily duration) lower stroke risk.

These findings were consistent after exclusion of smokers, and across exerciser and occupational status. One possibility is that these findings are reflective of short vigorous activity bursts contributing to balanced haemostasis, which can protect against haemorrhagic injury and mitigate arterial thrombosis, reducing the likelihood of stroke^36–38^. Further, these short bursts of activity may contribute to improved endothelial function^35^; this is particularly important for adults with treatment-resistance hypertension, where vigorous activity can reduce systolic blood pressure by up to 10 mm Hg^39^.

Higher stroke risk from long vigorous intensity bouts could stem, in part, from regular prolonged activity bouts inducing adverse vascular stress leading to adrenal gland dysfunction and subsequently dysregulated blood pressure. In addition, high shear vascular stress that can damage arterial walls leading to increased stroke incidence^10^ ^40^. When these repeated long activity bouts are combined with the pathophysiological effects of hypertension that induces increased arterial pressure and vascular damage, the risk of stroke may increase linearly. Collectively, our findings show the critical role of taking into account activity bout length in research among people with increased cardiovascular risk. Specifically, short bouts of high intensity activity may offer protective benefits against stroke, whereas prolonged bouts are a contributor to increased risk. If confirmed to be causal in future clinical trials, this could mark a significant refinement in how physical activity is prescribed for stroke prevention in adults with hypertension.

### Myocardial infarction and heart failure

We found short bouts of vigorous intensity and short/long bouts of moderate intensity were associated with lower risk for both myocardial infarction and heart failure. Long vigorous bouts also showed a protective association for myocardial infarction, albeit with wide 95% CI’s, whereas there was no protective association for heart failure.

This divergence in association patterns between myocardial infarction and heart failure for long vigorous intensity bouts may reflect the distinct pathophysiological mechanisms underlying these two conditions. Specifically, prolonged vigorous intensity bouts may lead to excessive strain on the cardiac and vascular systems, potentially leading to left ventricle related systolic or diastolic heart failure^41^ ^42^. In the case of myocardial infarction, long vigorous intensity bouts can reduce atherosclerotic plaque and mitigate the progression towards thrombotic or coronary occlusion^43–46^.

There can be possible contraindications of engaging in repeated sustained vigorous intensity activity, such as increased risk of induced cardiac ischemia and arrhythmias^47^. This may explain why short bouts of vigorous intensity had a stronger magnitude of association and tighter confidence intervals for myocardial infarction compared to long bouts of vigorous intensity. Our findings, alongside recent device-based studies focused on short bouts of high intensity activities^19^ ^20^ ^48^ ^49^, provide valuable insights for potentially reevaluating treatment options for patients with hypertension. Our results show conventional approaches that only consider total activity volume can mask the potentially health-enhancing benefits of short vigorous activity bouts and the adverse effects of repeated sustained and prolonged activity bouts. Consideration of how activity is accumulated can be clinically important, beyond total volume, in prevention of cardiovascular events for adults with hypertension.

### Strengths and limitations

Our sample was selected using a combined clinical definition of hypertension through linked hospitalisation, general practitioner, and medication records. The wearable devices, capable of capturing continuous and objective movement, in combination with the machine learning-based schema allowed us to measure physical activity at a high resolution to differentiate short and long bouts, which had not previously been possible with self-report questionnaires. Our results were robust to alternative bout thresholds, the exclusion of smokers, and participants with high frailty, fair to poor self-rated health, an event in the first five years of follow up, and pre-existing CVD. Despite these extensive precautionary measures, the potential for reverse causation caused by prodromal disease may still exist. Due to the observational design, we cannot rule out the presence of unmeasured or residual confounding. However, our E-values suggest an unmeasured confounder would need to have a strong association between 2.45 to 2.90 with the exposure and outcome for the relationship to be null. There was a median lag of 5.5 years between the UK Biobank baseline when covariate measurements were taken and the accelerometry study, although covariates were generally stable over time, except for medication.

## Conclusion

Among adults with hypertension, lower risk for MACE and MACE subtypes may be achieved through short bouts of vigorous intensity activity over moderate intensity alone. We found long bouts of vigorous intensity were associated with a steep 2-3 fold higher stroke incidence and generally was not associated with lower risk for other cardiovascular outcomes. Our findings underscore the need to reconsider the evidence base particularly with respect to vigorous intensity activity, and provide critical information for optimising personalised physical activity prescription and guidelines for CVD prevention in people with hypertension.

## Data Availability

All data produced in the present work are contained in the manuscript

## Figure titles and legends

**Figure 1:**

**Title:** Dose-response association of short and long moderate and vigorous intensity bouts with major adverse cardiovascular events

**Legend:** adjusted for age, sex, smoking status, alcohol consumption, light intensity, sedentary time, sleep duration, non-exposure intensity duration (eg: vigorous intensity, and >3 minute bouts of moderate intensity for up to 3 minute moderate intensity analysis), diet, prevalent cancer, parental history of CVD/cancer, education, frailty, and medication use

**Figure 2:**

**Title:** Dose-response association of short and long moderate and vigorous intensity bouts with stroke incidence

**Legend:** adjusted for age, sex, smoking status, alcohol consumption, light intensity, sedentary time, sleep duration, non-exposure intensity duration (eg: vigorous intensity, and >3 minute bouts of moderate intensity for up to 3 minute moderate intensity analysis), diet, prevalent cancer, parental history of CVD/cancer, education, frailty, and medication use

**Figure 3:**

**Title:** Dose-response association of short and long vigorous intensity bouts of varying lengths with stroke incidence

**Legend:** adjusted for age, sex, smoking status, alcohol consumption, light intensity, sedentary time, sleep duration, moderate intensity, non-exposure intensity duration (eg: >3 minute bouts of vigorous intensity for up to 3 minute vigorous intensity analysis), diet, prevalent cancer, parental history of CVD/cancer, education, frailty, and medication use

**Figure 4:**

**Title:** Dose-response association of short and long moderate and vigorous intensity bouts with myocardial infarction incidence

**Legend:** adjusted for age, sex, smoking status, alcohol consumption, light intensity, sedentary time, sleep duration, non-exposure intensity duration (eg: vigorous intensity, and >3 minute bouts of moderate intensity for up to 3 minute moderate intensity analysis), diet, prevalent cancer, parental history of CVD/cancer, education, frailty, and medication use

**Figure 5:**

**Title:** Dose-response association of short and long moderate and vigorous intensity bouts with heart failure incidence

**Legend:** adjusted for age, sex, smoking status, alcohol consumption, light intensity, sedentary time, sleep duration, non-exposure intensity duration (eg: vigorous intensity, and >3 minute bouts of moderate intensity for up to 3 minute moderate intensity analysis), diet, prevalent cancer, parental history of CVD/cancer, education, frailty, and medication use

